# SIR-PID: A Proportional-Integral-Derivative Controller for COVID-19 Outbreak Containment

**DOI:** 10.1101/2020.05.30.20117556

**Authors:** Aldo Ianni, Nicola Rossi

## Abstract

Ongoing social restrictions, as distancing and lockdown, adopted by many countries for contrasting the COVID-19 epidemic spread, try to find a trade-off between induced economic crisis, healthcare system collapse and costs in terms of human lives. Applying and removing restrictions on a system with uncontrollable inertia, as represented by an epidemic outbreak, may create critical instabilities, overshoots and strong oscillations of infected people around the desirable set-point, defined as the maximum number of hospitalizations acceptable by a given healthcare system. A good understanding of the system reaction to a change of the input control variable can be reasonably achieved using a proportional-integral-derivative controller, widely used in technological applications. In this paper we make use of this basic control theory for understanding the reaction of COVID-19 propagation to social restrictions and for exploiting a very known technology to reduce the epidemic damages through the correct tuning of the containment policy.

## 1 Introduction

The velocity of the Coronavirus disease outbreak (COVID-19), caused by severe acute respiratory syndrome coronavirus 2 (SARS-CoV-2) [1], is described, as for typical epidemics, by the so-called basic reproduction number (or ratio), usually denoted *R*_0_ [2] and defined as the expected number of new infections from a single new case in a population where all subjects are susceptible. Since a sizeable fraction of people infected by COVID-19, especially older and with underlying preexisting medical problems, are likely do develop serious illness with a fatality rate well above the typical seasonal influenza [3], many countries have adopted drastic or moderated social restrictions with the goal of reducing and controlling *R*_0_ by limiting the population mobility that determines the interaction among people.

From one side, drastic social restrictions, as the so-called lockdown, can not last too many weeks because it may cause serious damage to the economy; on the other side, relaxing too much the epidemic containment, in the so-called Phase-II or Phase-III may cause a restart of the pandemic growth setting a country in a status even more delicate. The best condition, would then be a compromise, in which *R*_0_ is brought close to the unity (or, better, ≤ 1) with the number of active cases reasonably close to a given threshold represented by the capability of a healthcare system to maximize the number of recovered people and to avoid the system collapse. This threshold is of course close to the maximum number of available intensive care hospitalizations. Hopefully this steady condition should be optimal while waiting for the realization and the commercialization of a vaccine, unfortunately not yet available for COVID-19.

The problem of manipulating the people mobility, and so trying to change *R*_0_, is in general not straightforward for huge inertia of epidemic propagation, because of the long incubation period (up to 14 days [4]) and the long recovery time (even close or larger than one month). In simple terms, it is not easy to act on a system with such a long latency and fast reaction to relaxation. This is the typical cases in many technological systems in which a control system tries to reach a set-point in a smooth way avoiding dangerous stress and unstable oscillations of the system itself. As a typical example one can think of maneuvers of a big ship or of speed control actuated by cruisers installed on modern cars.

The control system theory is quite complicated and different solutions have been studied over the centuries. One basic example, sometimes limited, but in general with high performance, is the proportional–integral–derivative controller (PID). In this paper we will try to apply the most important features of such a mechanism for understating the evolution of the COVID-19 outbreak spread when controlled with social restrictions. This modelling could help in understanding the present situation of many countries, and its description can be used to choose the right sequence of the social restriction sequence.

In Sec. 2 we will introduce the PID controller theory with its most important features. In Sec. 3 we will describe a mathematical model for the COVID-19 epidemic evolution based on time-dependent modification of SIR compartmental model. In Sec. 4 we will describe the numerical implementation of the resulting SIR-PID model. In Sec. 5 we will describe the tuning of the SIRPID coefficients. Finally in Sec. 6 we will recommend a simple way of implementing the mechanism on real data affected by large statistical uncertainty.

## 2 The PID Controller

A PID controller is a feedback control loop mechanism widely used in technical systems in which a continuous modulated control is required. Even if some basic properties were grasped a few centuries ago, the first mathematical formalization dates back to early 1900 [5]. Nowadays PID controllers are used in many automatic processes requiring high stability and optimization.

The PID computes on line the *error value* input *e*(*t*) as the difference between a desired set-point(SP) and a measured process variable (PV), received as feedback. A PID applies correction through the *control variable* output *u*(*t*) consisting of a weighted average of a proportional (P), integral (I) and derivative (D) term, according to the flow diagram reported in Fig. 1. The controller basically aims at minimizing the error over time by updating the control variable to a newer value established by control terms. Its explicit mathematical formulation reads:

**Figure 1:**
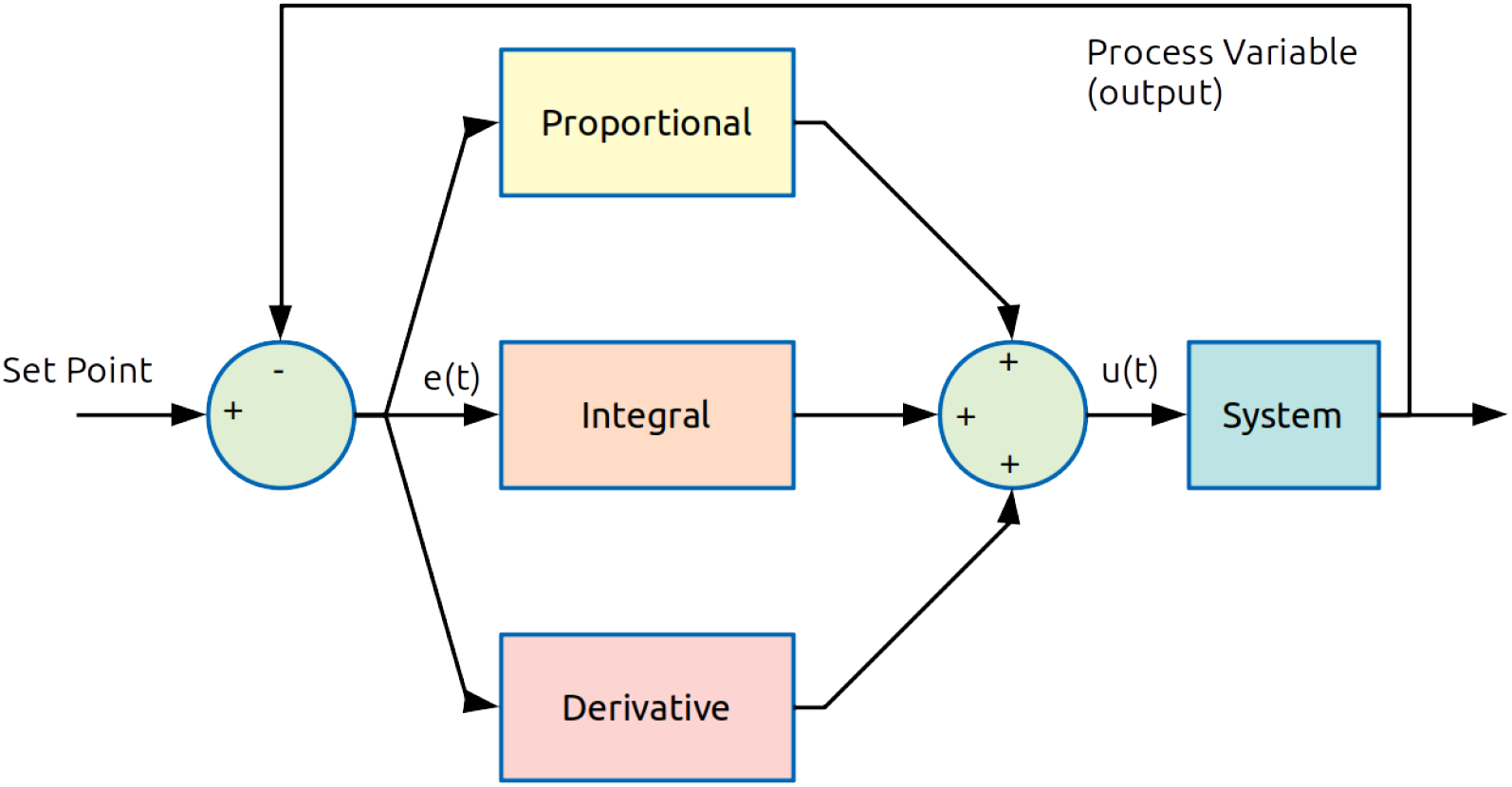
Flow diagram of the PID controller. The error function *e*(*t*) is the difference between SP and PV. The weighted average of P, I and D contributions determines the output control *u*(*t*) for the specific system to be controlled.

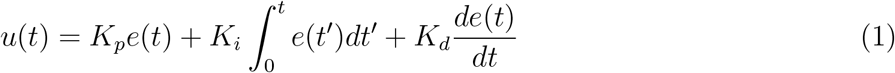

where *K_p_*, *K_i_* and *K_d_* represent the proportional, integral and derivative terms respectively.

The meaning of the three terms can be summarized as follows:

- **Proportional Term**: represents the action of the *present* condition of the system and it is proportional to *e*(*t*) through the coefficient *K_p_*. The proportional control alone generates a response only if *e*(*t*) is different from zero. In other words, if the error is positive, the control output is correspondingly positive and vice versa.
- **Integral Term**: takes into account the *past*, by adjusting the control output with the cumulative residual error *e*(*t*). The integral term stops growing when the set-point is reached, keeping the desired set-point.
- **Derivative Term**: gives an estimation of the *future* condition. The control is proportional to the error change rate. Fast change induces fast control and vice versa.

The optimized choice of the *K*’s parameters is achieved after a *loop tuning* process. Their value are related to the system response and they can be tuned with different methods observing the behaviour of the system response after fixing or moving a given set-point. For further detail see Sec.

## 3 The SIR-PID Model

In order to model the behavior of the COVID-19 epidemic applying social restrictions aiming at changing the basic reproduction number *R*_0_, we consider a basic SIR compartmental model [6], modified in order to account for a time dependent *R*_0_(*t*) as described in [7]. Hereafter we will refer to this model as SIR-PID. It is worth nothing that if the system response is known *a priori*, the control condition can be determined mathematically independently of the PID mechanism, or alternatively one can calculate the PID coefficients *a priori*. We will forget anyway this aspect and we will proceed as if the epidemic system response was completely unknown. This generalizes the specific control to a real epidemic system, as the COVID-19, i.e. not completely modelled in all its, sometimes unknown, details. In the model we distinguish three categories of individuals, known as *compartments*: *S*(*t*)
for the number of susceptible, *I*(*t*) for the number of infectious and *R*(*t*) for the number of recovered, or better *removed*. The last category might include recovered and deceased individuals together or separately. Based on these assumptions the mathematical model is written in terms of the following set of differential equations:

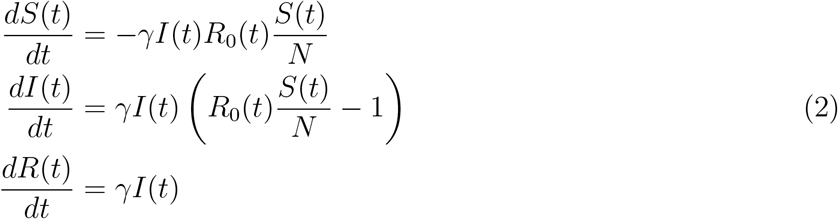

where *N* = *S*(*t*) + *I*(*t*) + *R*(*t*) is constant by definition being zero the sum of their derivatives and *γ* is the *recovery rate*. In order to match the standard SIR model *γR*_0_ corresponds to *β*, usually called *transition rate*.

If a susceptible population of (*S*(*t* = 0) = *N*) people is affected for the first time by the disease outbreak, the initial conditions of the system (2) can be easily cast as: *N*(0) = *N*_0_, *I*(0) = *I*_0_ and *R*(0) = 0. In the basic SIR model, as reported above, *R*_0_ determines the dynamic of the infection, and emerges as the ratio *β/γ*. With this definition we focus on the second equation in (2):

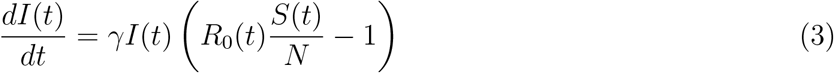

From eq. (3) it follows: 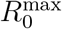 Therefore, the epidemic decreases when *R*_0_ <*N/S*(0).

Figure 2 shows the evolution of the compartments in case we assume that the basic reproduction number decreases exponentially as a function of time. This represents a typical situation when social restrictions are applied progressively from mild to strong conditions, see [7] for further details.

**Figure 2:**
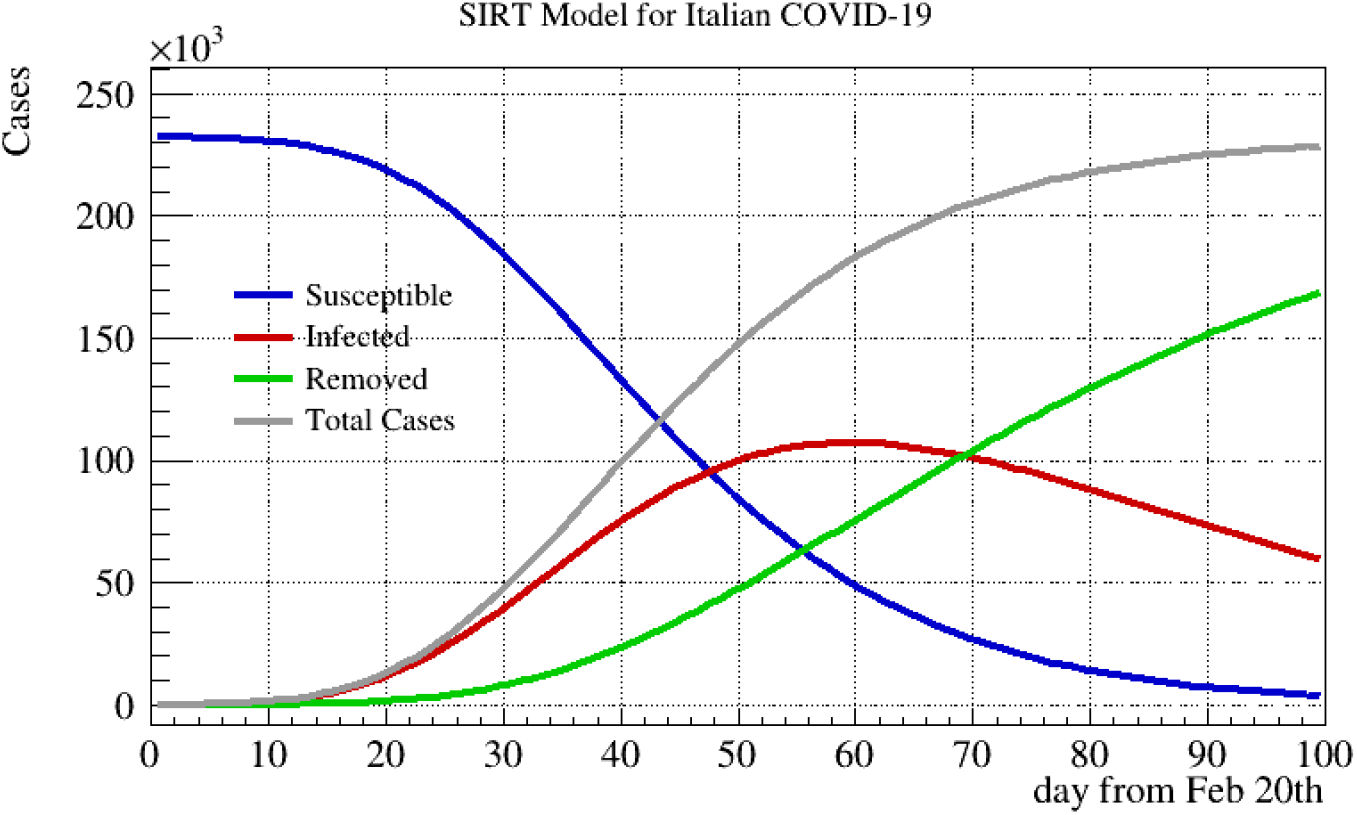
Example of time dependent SIR model in which the basic reproduction number decreases exponentially since the beginning of the disease epidemic outbreak. The initial susceptible population S (blue) is converted into infected I (red) according to the time-dependent *R*_0_. Finally, according the strength of *γ*, I are converted into removed R (green). The grey curve represents the sum R+I, and usually is very well approximated by a logistic curve.

## 4 SIR-PID Numerical Implementation

In order to implement and study a simple SIR-PID model, let us group the standard compartment functions S(t), I(t) and R(t) in a vector 𝕐 = (*S, I, R*). The system (2) can be easily recast as

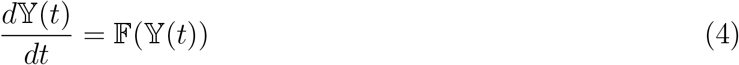

with initial condition: 𝕐(0) = 𝕐_0_, where 𝔽 represents the non-linear vectorial function defined in the SIR equations.

The system can be solved numerically using e.g the Runge-Kutta method [8] even if, for the sake of simplicity, we will consider a simple forward Euler’s method [9]. The latter states that for each constant iteration Δ*t* = *t_n_*_+1_ *− t_n_*, the solution of the differential equation can be incremented as:

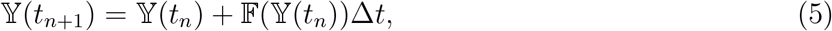

In particular, the second component of 𝕐(*t*) representing the infected people or active cases

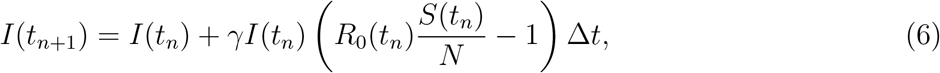

is the output of our PID controller, while *u*(*t*) = *R*_0_(*t*)/*A_R_* is the input control variable, where *A_R_* is a normalization constant, that can be chosen equal to one for having 100% at *R*_0_ = 1. Defining the error *e*(*t_n_*) = (*I*_SP_ − *I*(*t_n_*))/*A_I_* (with e.g. *A_I_* = *I_SP_* for normalizing the response), the SIR-PID model is then implemented as

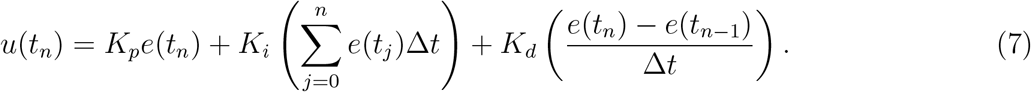

The second and third term of the right-hand side represent the numerical approximation of the integral and the derivative of the error *e*(*t*), respectively. Finally, let us put some reasonable ingredients in our model:

- Let *N* = 10^7^be the typical population of a small country or region. In order to understand the SIR-PID response, N must be greater then the typical infected population, in order to avoid the so-called *herd immunity* turning point as expected for free evolution of epidemics without any attempt of containment.
- We can imagine that the outbreak starts with an out-of-control *patient zero*. The initial condition is therefore: 𝕐 = (*N*, 1, 0).
- *R*_0_(*t*) can range from 0.5 to 10. We assume that a country cannot do a total lockdown (*R*_0_ = 0), being some basic goods and services still guaranteed. *R*_0_ = 10 represents in this case the maximum reproduction number, corresponding to absence of actions by governments and/or by freedom of action by citizens. This value is assumed as starting condition for the control variable.
- Social restrictions are applied by decrees every two weeks (Δ*T* = 14 days) and, after application, *R*_0_ decreases anyway by inertia of one unity per day.
- Let set-point be *I*_SP_ = 10000, representing a threshold chosen in order not to make the health-care system collapse and guarantee a certain number of intensive cares.
- Let us choose *γ* = 1*/*30 assuming one month for the typical recovery time.
- In order to guarantee a good accuracy of the solutions of (4) with the forward Euler’s method, let set Δ*t* = 0.0001 day.

The system is free to evolve and ready for tuning.

## 5 Tuning and Interpretation

The tuning process aims at the best calibration of the proportional gain, the integral gain and the derivative correction selecting the proper proportional bands. The final goal is to reach the system stability within the desired range with a desired time scale. The PID tuning is a quite complex problem because each system has a different (very often unknown) response, even if the selection of the P, I, and D parameters may look quite intuitive.

Many different procedures have been developed over years. Some empirical rules, as the Ziegler-Nichols tuning [10] looks quite flexible and usable for the majority of the technological applications. It is also possible to determine the PID coefficients performing complex system simulations and there are a lot of algorithms of self-tuning. For further details, see e.g. [11].

In Fig. 3 we show a generic system responding respectively to a pure proportional control (red), a proportional corrected with the integral term (yellow) for removing the bias with the set-point (dashed black) and finally with oscillation damped by the integral term (violet).

**Figure 3:**
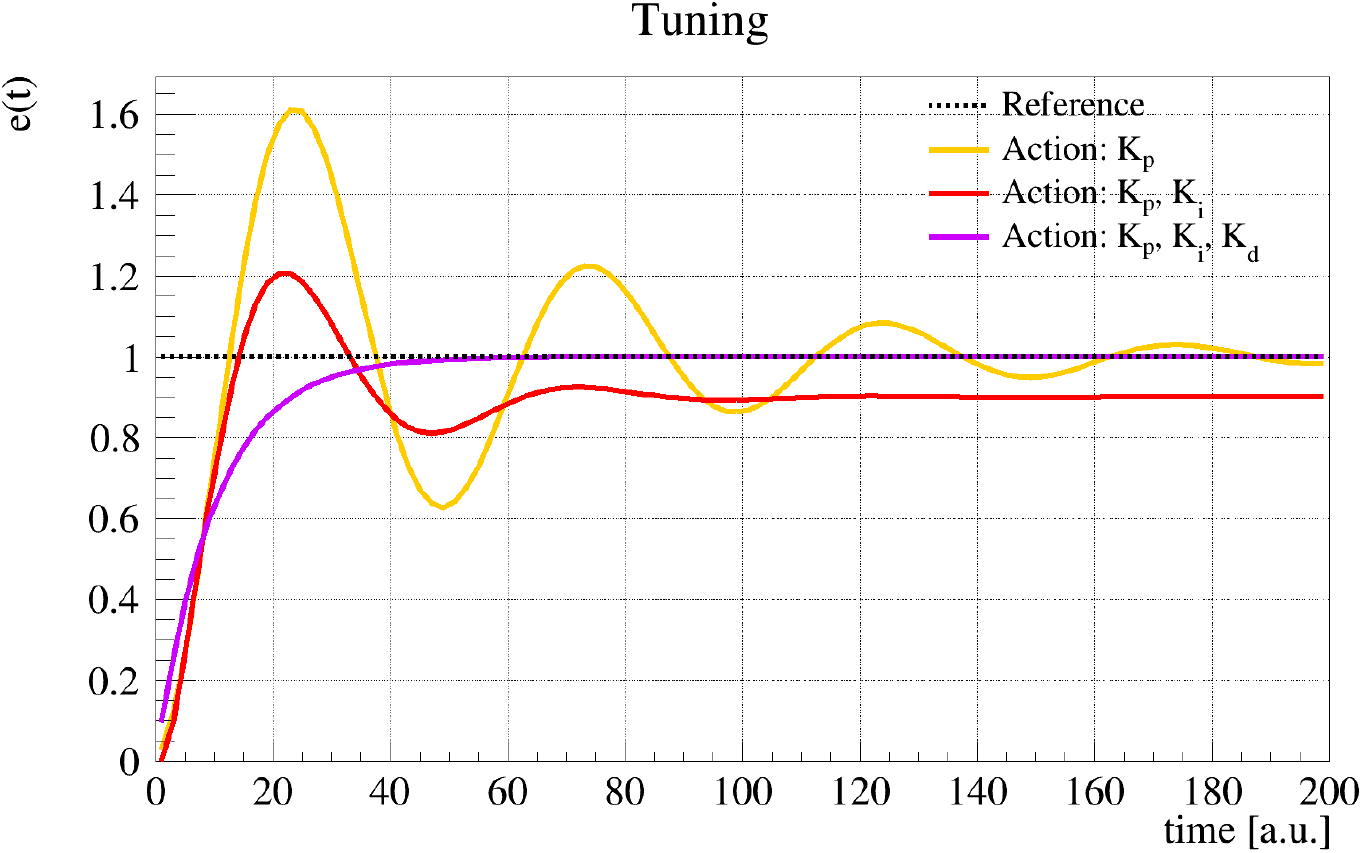
Tuning examples: a generic system responding respectively to a pure proportional control (red), a proportional corrected with the integral term (yellow) for removing the bias with the set-point (dashed black) and finally with oscillation damped by the integral term (violet).

Following a manual tuning we will report different behaviors of the SIR-PID introducing step by step different values for the PID coefficients.

Figure 4 shows behavior of input *R*_0_ (top) and output *I*(*t*) bottom with *K_p_* = 10, *K_i_* = *K_d_* = 0. The dashed black line represents the set-point. A very strong proportional action only is not able to stabilize the system. This corresponds to complete absence of restrictions at the beginning and a drastic lockdown when the threshold is reached, and this behavior is repeated every time the set-point is reached, causing many out-of-control devastating epidemic waves. This scenario could happen when an excessive optimism makes the restrictions relax too early until the situation becomes again out-of-control and another drastic lockdown is needed.

**Figure 4:**
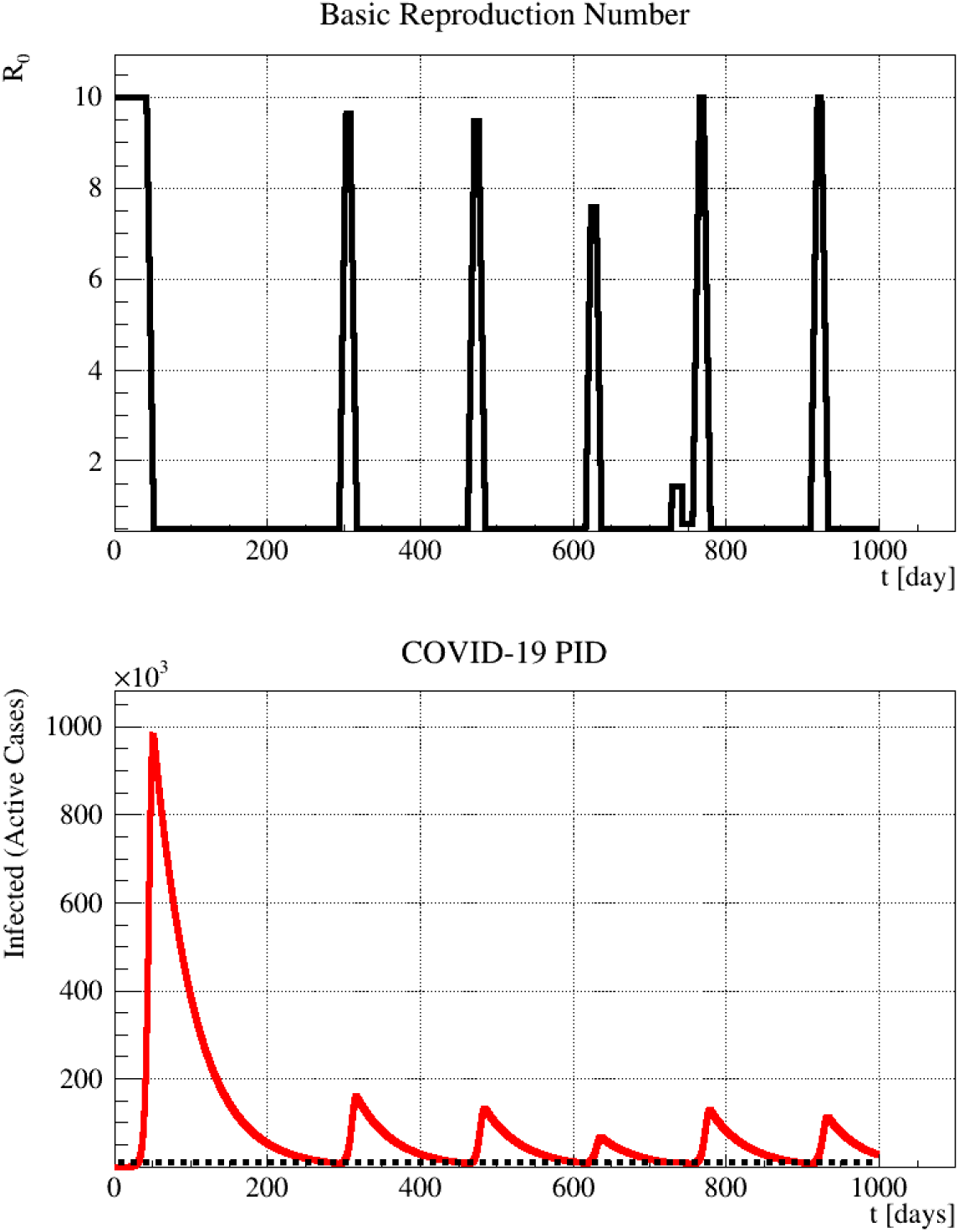
Behaviour of input *R*_0_ (top) and output *I*(*t*) bottom with *K_p_* = 10, *K_i_* = *K_d_* = 0. The dashed black line represents the set-point. A very strong proportional action only is not able to stabilize the system. This corresponds to complete absence of restrictions at the beginning and a drastic lockdown when the threshold is reached, and this behavior is repeated every time the set-point is reached, causing many out-of-control devastating epidemic waves.

In Fig. 5 we show the behavior of input *R*_0_ (top) and output *I*(*t*) (bottom) with *K_p_* = 5, *K_i_* = *K_d_* = 0. A milder proportional term avoids a strong overshoot at the first reach of the set-point but it is not sufficient for damping the system oscillation. This corresponds to a less drastic social restriction at the beginning and then a subsequent attempt to stabilize the rate with a mild and strong lockdown. This situation could be in principle acceptable, yet the residual oscillations can create instabilities and make people unhappy with continuous change of the restrictions policy.

**Figure 5:**
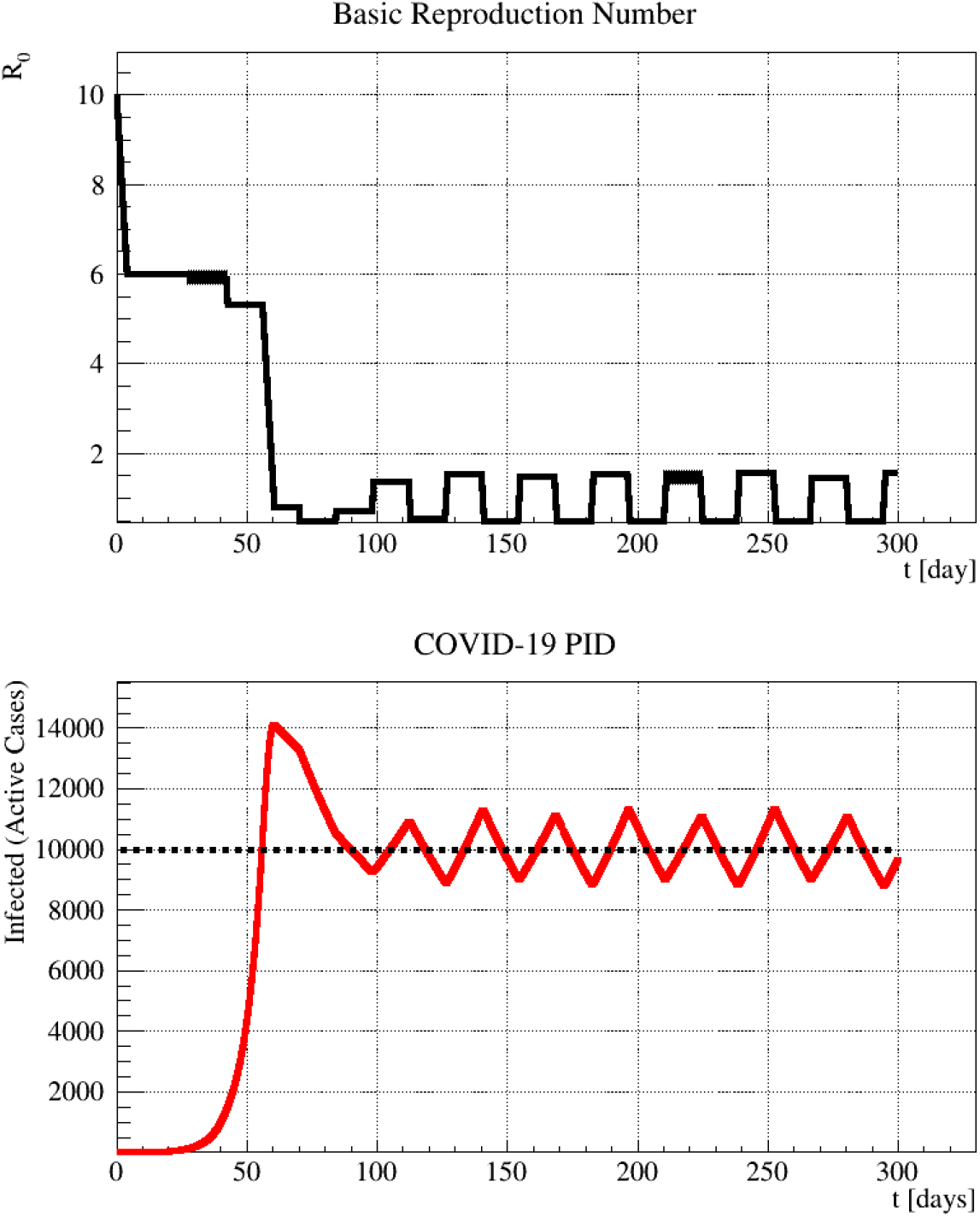
Behaviour of input *R*_0_ (top) and output *I*(*t*) bottom with 5 and *K_i_* = *K_d_* = 0. The dashed black line represents the set-point. A mild proportional term avoids a strong overshoot at the first reach of the set-point but it is not sufficient for damping the system oscillation. This corresponds to a less drastic social restriction at the beginning and then a subsequent attempt to stabilize the rate with mild and strong lockdown.

Finally, in Fig. 6 we show the behavior of input *R*_0_ (top) and output *I*(*t*) (bottom) with optimal tuning *K_p_* = 2.4, *K_i_* = 0.04 and *K_d_* = 0.004. The optimal tuning, that includes the integral and derivative term, allows to reach the set-point smoothly. This corresponds to a mild lockdown at the beginning and a subsequent fine tuning of the restriction, corresponding to the final social distancing that can remain unchanged. This scenario is optimal and can be afforded by a country in order to avoid strong economic damages, finding a good compromise while waiting for a different solution, as vaccination.

Even this last scenario could need further corrections because of many other effects, like the possible change of *R*_0_ due to the environmental conditions or to possible attenuation of the COVID-19 virulence. That is the reason why we warned at the beginning that the SIR-PID model is only a test-bench theoretical model and the actual response must be found probing the real COVID-19 system with PID control actions.

**Figure 6:**
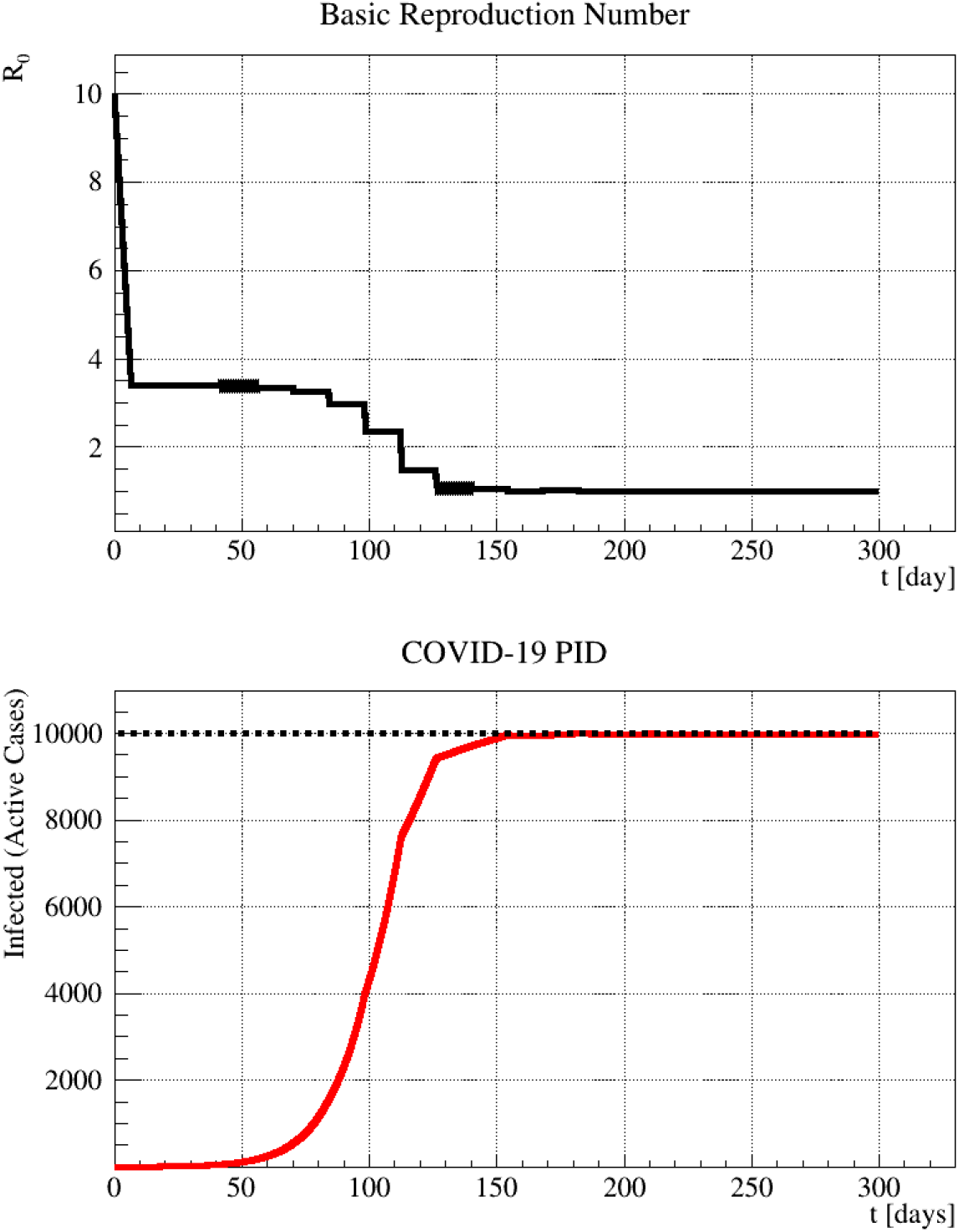
Behavior of input *R*_0_ (top) and output *I*(*t*) (bottom) with tuned *K_p_* = 2.4, *K_i_* = 0.04 and *K_d_* = 0.004. The dashed black line represents the set-point. The optimal tuning, that includes the integral and derivative term, allows to reach the set-point smoothly. This corresponds to a mild lockdown at the beginning and a subsequent fine tuning of the restriction, corresponding to the final social distancing that can remain unchanged.

## 6 Application on Epidemiological Data-sets

Working with real data it is not that straightforward because of large fluctuations on sampled data, basically due to the intrinsic statistical nature of the phenomenon and, more important, due to the discontinuous procedure of administration of medical tests. For that reason the computation, especially of the derivative, can be affected by large uncertainty that may cause a false understanding of the system reaction.

It is a good practice therefore to smooth the data using for example a local regression (LOESS) method based of Savitzky-Golay filter [13]. For instance, Fig. 7 shows the application of the local regression (red curve) on Italian daily new cases (orange dots) [12]. Once the data are flattened by the smoothing process, the integral can be numerically computed using e.g. the Simpson’s rule [15], while a stable derivative estimation can be performed applying the Richardson’s extrapolation method [14].

**Figure 7:**
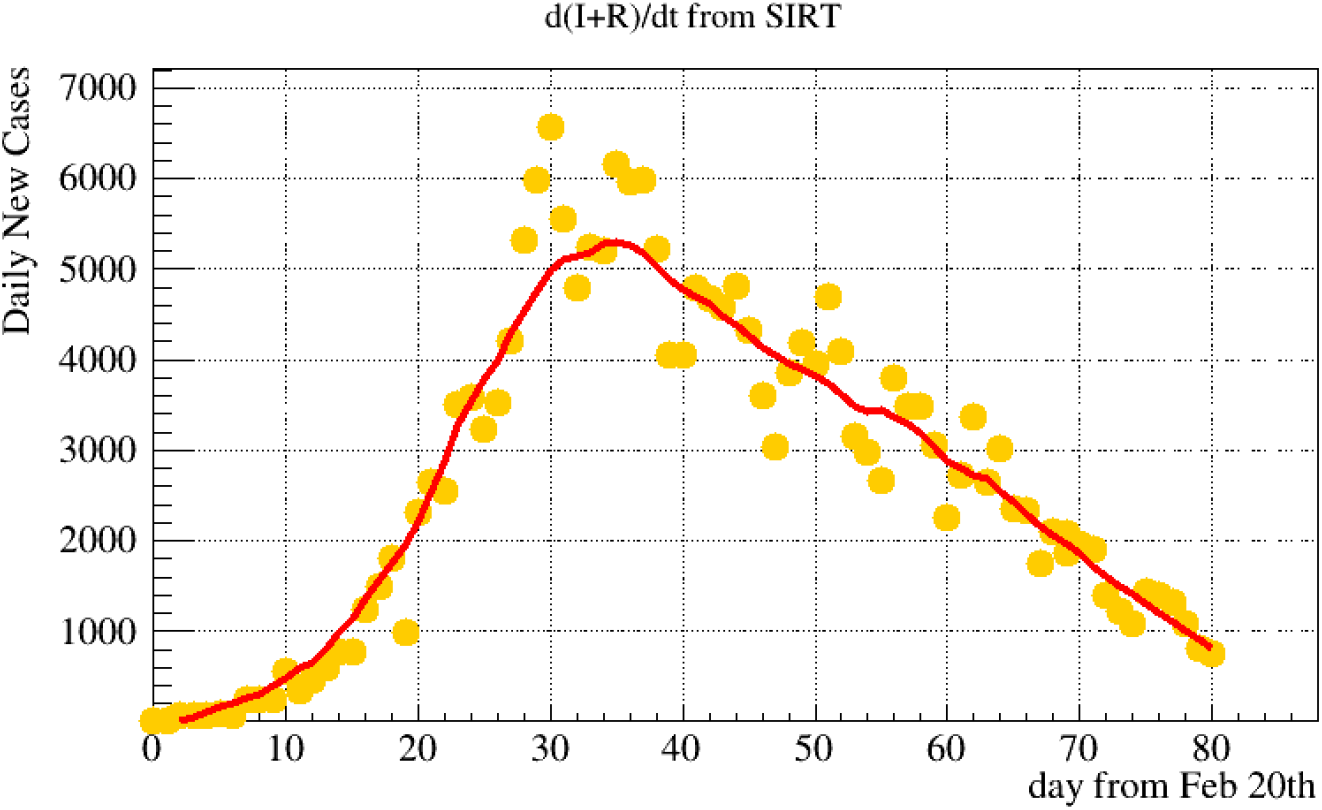
Daily new cases in Italy (yellow dots) since the beginning of the COVID-19 outbreak and application of the LOESS filter (red curve).

We are considering *R*_0_ as input parameter even tough it is not generally easy to determine this parameter from the data with an ongoing outbreak, see again [2]. As a consequence, *R*_0_ has to be interpreted, at leading order, in terms of people mobility even if environmental effects and intrinsic changes can modify the COVID-19 virulence. If we make this simple hypothesis, *R*_0_, from 0 to 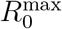 (10, in our example), can be interpreted as 10 levels of mobility, from drastic lockdown (*R*_0_ = 0) to free epidemic evolution 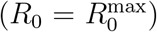. Values in between represent different gradations of the social distancing ruled by legislative decrees settles every 14 days, the typical time scale of the COVID-19 epidemic latency.

The mobility can be determined *a priory* considering the daily traffic of different society compartments as e.g. basic goods production/services, massive industrial production, shops and markets, public transportation, open-air sports, gyms and entertainments, etc… This mobility can include or not the usage of protections devices (as masks, gloves and disinfectant) and the application of safety distances, or limit number of indoor people, and so on. Once the mobility scale from 0 to 10 is defined according to mobility/protection estimations one can test the SIR-PID model on the real COVID-19. Probing the system one can study the optimal tuning of the SIR-PID coefficient in order to reach smoothly the desired set-point minimizing the outbreak damages.

## 7 Conclusions

In this paper we have shown that the COVID-19 outbreak spread with the attempt of containment through social restrictions by national governments, can be modelled and understood in terms of the PID controller mechanism, widely used on technical applications on complex systems.

Using a simple time-dependent modification of the SIR modelling of the COVID-19 outbreak evolution, we built a test-bench model called SIR-PID for testing the possibility of using a PID controller for achieving the desired containment threshold smoothly, aiming at avoiding serious damages in terms of economical crisis and especially in terms of human life costs. The implementation assumes the basic reproduction number as input parameter and the number of infected (or active cases) as output. Even if *R*_0_ is not directly accessible as input parameter, it can be reasonably substituted by the people mobility that can be easily estimated and classified in a gradation scale ranging from the drastic lockdown to the free evolution of the pandemic.

We showed that using a loop tuning procedure this goal is achievable in a quite satisfactory way. This result allow us to exploit this procedure on real data, even if the real COVID-19 outbreak system could be a bit more complex due to other potential effects contributing to the time variation of *R*_0_. For our basic model we show that the best way of achieving an optimal control is to react promptly at the beginning of the pandemic by lowering the mobility of a factor two and then increase the social restrictions slowly in order to reach the desired set-point, affordable by the healthcare system. Anyway this procedure is desirable when the pandemic is already out-of-control over a wide geographical area, because the first attempt should be of course the complete confinement of the outbreak, by drastically limiting the patient zero area and nearest contacts, when possible.

## Data Availability

https://www.worldometers.info/coronavirus/

## Acknowledgments

We would like to thank also Francesca Baldinelli for helping in understating some details about *R*_0_ and Guido Baldinelli for explanations about the medical meaning of different COVID-19 aspects. We also thanks all people contributing in proofreading, even with very short comments.

